# Development and Accuracy Determination of a Peptide Diagnostic Based on the N-terminal Ectodomain of the Membrane Glycoprotein

**DOI:** 10.64898/2026.07.04.26355775

**Authors:** Brian Andrich L. Pollo, Julia Patricia B. Llagas, Riziel Hannah B. Aguimatang, Ayra Patrice N. Espiritu, Danica S. Ching, Maria Isabel C. Idolor, Ruby Anne N. King, Fresthel Monica M. Climacosa, Salvador Eugenio C. Caoili

**Affiliations:** Biomedical Innovations Research for Translational Health Science (BIRTHS) Laboratory, College of Medicine, University of the Philippines Manila, Manila, Philippines; Institute of Clinical Epidemiology, National Institutes of Health, Manila, Philippines; Department of Science and Technology – Philippine Council for Health Research and Development (DOST-PCHRD), Taguig, Philippines; Department of Medical Microbiology, College of Public Health, University of the Philippines Manila, Manila, Philippines

**Keywords:** Peptides, Antibodies, COVID-19, Enzyme-Linked Immunosorbent Assay, Protein Binding

## Abstract

**Background:** The N-terminal ectodomain (NTE) of the SARS-CoV-2 membrane (M) glycoprotein is a short, flexible region that remains exposed on the virion surface and exhibits immunogenic potential across multiple coronaviruses. Despite its small size and conformational plasticity, this region contains conserved linear epitopes that may serve as practical surrogates for full-length proteins in serological diagnostics.

**Objective:** To develop and evaluate a synthetic peptide-based diagnostic assay targeting the NTE of the SARS-CoV-2 M protein.

**Methods:** Epitope prediction, peptide synthesis, and antibody affinity assays were performed to design homomultivalent peptide analogs that exploit avidity effects through disulfide polymerization. The resulting peptide antigens were tested in an enzyme-linked immunosorbent assay (ELISA) using clinical samples from RT-PCR–confirmed COVID-19 patients and biobanked controls.

**Results:** The selected peptide analogs (M1, M1i, M1s) corresponded to a conserved surface-exposed motif of the SARS-CoV-2 M protein. Polymeric M1 exhibited a twofold gain in apparent affinity (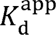 = 4.33 nM) compared with the monomeric form (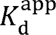 = 8.00 nM). Clinical validation using 1,222 patient samples yielded a sensitivity of 95.26% and specificity of 52.27%, with an overall diagnostic accuracy of 88.70%.

**Conclusion:** The M peptide analogs demonstrate that synthetic peptide antigens can serve as stable, high-sensitivity surrogates for whole-protein assays. This design principle may be applied to other emerging pathogens where rapid assay development and scalability are critical.

## Introduction

Since the emergence of SARS-CoV-2 in late 2019, the rapid and scalable development of diagnostic tools has been crucial for global public health response [1]. Although most serological assays target large recombinant proteins such as the spike (S) or nucleocapsid (N), these require complex expression systems, purification pipelines, and cold-chain logistics [2,3]. Peptide-based diagnostics, in contrast, offer significant advantages in synthesis speed, stability, and modularity [4,5]. However, their primary limitation lies in reduced antibody-binding affinity, which can compromise sensitivity [6].

The membrane (M) glycoprotein, while less frequently studied than S or N, plays a central role in virion assembly and host immune recognition [7]. Its N-terminal ectodomain (NTE) extends outside the viral envelope and exhibits flexible conformation conducive to B-cell recognition [8]. Sequence analyses of coronaviruses indicate that the M NTE harbors conserved, surface-accessible epitopes across SARS-CoV, MERS-CoV, and SARS-CoV-2, suggesting potential for diagnostic or vaccine applications [9].

Previous work has focused primarily on peptide epitopes derived from the spike protein, with mixed results in clinical sensitivity [10]. The NTE, being shorter and more hydrophilic, may yield peptides that mimic antigenic surfaces more effectively when presented in multivalent forms [11]. Polymerization of peptides through disulfide linkages can simulate epitope clustering, enhancing apparent antibody affinity through avidity [12,13].

This study aimed to design synthetic peptide analogs of the SARS-CoV-2 M NTE, characterize their antibody-binding affinity, and evaluate the diagnostic performance of an ELISA incorporating these analogs using human clinical samples collected prior to vaccination rollout.

## Materials and Methods

### Design of synthetic peptides

A synthetic peptide library targeting the SARS-CoV-2 spike protein was developed to identify potential immunogenic epitopes. The design process involved querying the Immune Epitope Database for linear B-cell epitopes with confirmed positive assay results from SARS-CoV-2. The retrieved sequences underwent cluster analysis to group similar epitopes. Structural and functional analyses were conducted by retrieving three-dimensional structures from the Protein Data Bank (PDB; https://www.rcsb.org/) and UniProt (https://www.uniprot.org/), visualized using UCSF ChimeraX (version 1.6.1; https://www.rbvi.ucsf.edu/chimerax/). Peptide sequences were selected based on the following criteria: predicted intrinsic disorder, length between 12 and 20 amino acids, absence of residues that could interfere with polymerization (*e.g.,* cysteine, methionine, tryptophan, lysine), and hydrophobicity less than 50%. Sequence homology was assessed using BLASTp to ensure specificity to SARS-CoV-2 epitopes. The selected peptide cores were modified by adding cysteine residues at the N- and C-termini to facilitate polymerization through disulfide bond formation.

### Synthesis and polymerization of peptides

The antigenic peptides were synthesized by a commercial provider specializing in peptide synthesis. Solid-phase peptide synthesis was employed, and the peptides were analyzed for purity and identity using high-performance liquid chromatography (HPLC). The peptides were then subjected to polymerization under mild oxidizing conditions. Dimethyl sulfoxide (DMSO) was added to achieve a final concentration of 50%, and the mixture was incubated at room temperature for 24 hours to promote disulfide bond formation.

### Preparation of antipeptide antibodies (ApAbs)

New Zealand White rabbits were immunized with keyhole limpet hemocyanin (KLH)-conjugated M1i peptides emulsified in Freund’s adjuvant (Day 0, 14, 28). Sera were collected on Day 42 and affinity-purified on M1-Sepharose 4B columns.

### Measurement of Avidity Gain by Depolymerization using Compositionally Matched Oxidation-Reduction with N-acetylcysteine (CORN)

The lead candidate peptide underwent depolymerization to assess the contribution of the polymeric structure to avidity. N-acetylcysteine (Nac) was used as a reducing agent to break disulfide linkages. Aliquots of the polymeric peptide were incubated with various concentrations of Nac at 37°C for 30 minutes. To match the composition of the test solutions with the negative control, DMSO was added to a final concentration of 20%. The negative control was prepared by heating Nac in the presence of DMSO to degrade it. The complete degradation of Nac was confirmed using a specific assay and by the characteristic odor of dimethylsulfide. The depolymerized peptide was then derivatized with trinitrobenzene sulfonate (TNBS) and analyzed using an enzyme-linked immunosorbent assay (ELISA) with anti-TNP as the primary antibody. The apparent dissociation constant (KDapp) was calculated for both the polymeric and depolymerized forms of the peptide.

### Affinity determination using ApAbs

To assess peptide-antibody interactions, indirect ELISA was performed as previously described in protocols.io [14]. Peptides were diluted in coating buffer to achieve a final concentration of 10 nM peptide and used to coat high-binding 96-well microtiter plates. The plates were incubated at 4°C overnight, followed by washing with phosphate-buffered saline containing 0.05% Tween-20 (PBST). Derivatization was carried out by adding TNBS solution and incubating at 37°C for 15 minutes. Unreacted TNBS was quenched with glycine, and the plates were washed again. Blocking was performed with 2% skimmed milk in PBST at 37°C for 30 minutes. Serial dilutions of anti-TNP antibody were then applied, followed by incubation with protein A peroxidase conjugate. Chromogenic development was achieved using tetramethylbenzidine and hydrogen peroxide, and the reaction was halted with 0.1□M sulfuric acid. Absorbance was recorded at 450□nm, and KDapp values were calculated from the slope of linearized plots.

### Immunoassay Optimization

Selected M NTE analogs underwent optimization for immunoassay performance. ELISA was performed to confirm peptide immobilization on polystyrene plates using TNP-labeled peptides and anti-TNP antibody. Optimal antigen concentration was determined using pooled COVID-19 sera, with 20 μg/mL selected for subsequent testing on clinical samples. Assay performance metrics were assessed across a serial geometric dilution of human serum (1:100 to 1:6400). Indirect ELISA was performed without the depolymerization and derivatization steps to evaluate assay sensitivity and specificity.

### Ethical approval and patient enrollment

The study was conducted according to STARD guidelines for reporting diagnostic accuracy studies. Ethics approval was obtained from the University of the Philippines Manila Research Ethics Board (Code 2022-0492-01). The full protocol is registered under HERDIN (https://www.herdin.ph/) (PHRR230601-005772). All authors had access to study data and approved the final manuscript. Laboratory personnel performing ELISA were blinded to patient disease status throughout testing.

### ELISA optimization using collected samples

Candidate peptides demonstrating the strongest reactivity were subjected to further optimization. Immobilization efficiency was verified using TNBS-labeled peptides detected with anti-TNP antibody. Optimal coating concentration was established by testing pooled patient sera at varying antigen concentrations, with 20□μg/mL selected for subsequent testing. Serial geometric dilutions of serum (1:100 to 1:6400) were performed to assess assay sensitivity, specificity, and linear dynamic range. Indirect ELISA was performed without depolymerization or derivatization for all optimization assays.

### Patient recruitment and sample collection

Clinical samples were collected from patients hospitalized at a tertiary hospital between October 2020 and February 2021. Inclusion criteria included being at least 18 years old and having RT-PCR-confirmed COVID-19 from nasopharyngeal swab. Convenience sampling was utilized, with potentially eligible participants being referred by their physician-in-charge. RT-PCR testing was performed by hospital laboratory personnel following standard operating procedures. Serum samples were collected on days 1, 7, and 14 of hospitalization and stored at −80°C before use. Disease severity was classified by attending clinicians according to patient presentation on admission: mild (no pneumonia), moderate (with pneumonia), severe (requiring supplemental oxygen), or critical (requiring mechanical ventilation or signs of septic shock). Control serum samples were obtained from healthy blood donors prior to the emergence of SARS-CoV-2 in December 2019. These samples served as negative controls for all assays. All serum samples were heat-inactivated at 56°C for 30 minutes and stored at −80°C before use.

### Diagnostic Accuracy Analysis

Diagnostic accuracy measures were calculated using statistical software. The *post hoc* cutoff was determined based on Youden index. Measures were calculated based on both the prespecified threshold and the exploratory cutoff. Sample size calculation was performed using power analysis software, with an estimated prevalence of 42.3% and a null hypothesis that the ELISA would yield a binary result.

## Results

### Design and structural analysis of synthetic peptide analogs of the SARS-CoV-2 M NTE

Mapping onto crystal structures revealed that the M1 region corresponds to a disordered, unresolved N-terminal segment preceding the first transmembrane helix (residues 1–25). Despite the lack of defined secondary structure, this segment is predicted to extend outward and carry a net positive charge at physiological pH, favorable for antibody recognition. Structural visualization using UCSF ChimeraX (version 1.6.1) and crystallographic data from PDB entries 7VGR, 7VGS, and 8CTK confirmed that this NTE region remained largely unresolved in available M protein crystal structures (**Figure 1**). The unresolved residues, denoted by asterisks in the sequence map, corresponded to a conformationally flexible segment lacking well-defined secondary structure. Predicted α-helical elements began only after residue 10, as indicated by the helical notation (H). Both M1 (CADSNGTITVEELKKLLEQC) and its internal variant M1i (MADSNGTITVEELKKLLEQC) occupied surface-exposed positions, implying accessibility to antibodies.

**Figure 1.**
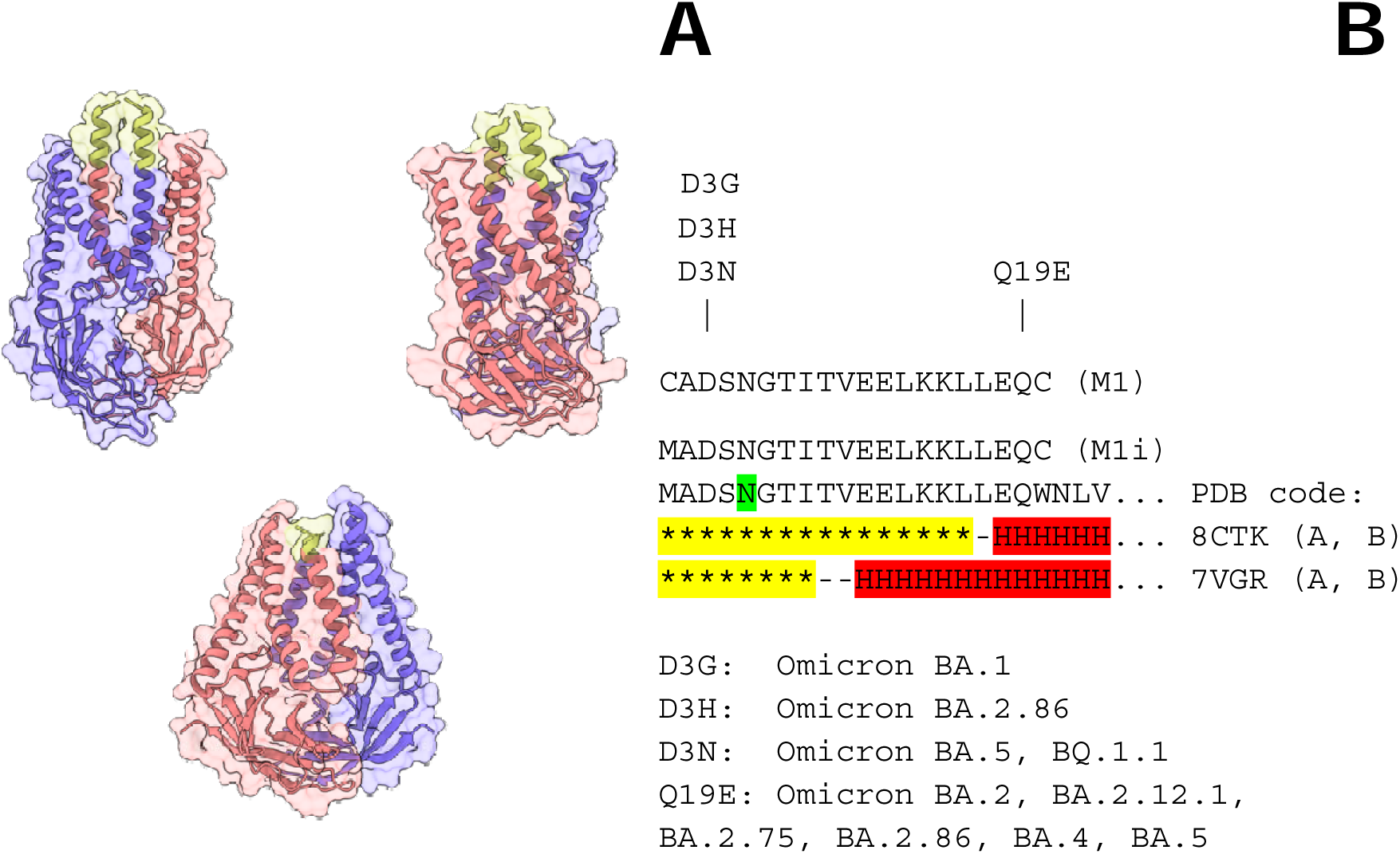
Structural and sequence characteristics of the SARS-CoV-2 M protein N-terminal ectodomain (NTE). (A) Three available M protein dimer structures were visualized using UCSF ChimeraX 1.6.1, highlighting the NTE region in yellow. PDB entries 7VGR (top left), 7VGS (top right), and 8CTK (bottom) represent distinct conformational states of the membrane-associated dimer [33]. The highlighted NTE segment corresponds to a surface-exposed, flexible domain at the periphery of the transmembrane interface, consistent with potential accessibility to host immune factors. (B) Sequence alignment of M1 and its variant M1i demonstrates that this portion of the NTE is highly conserved across major SARS-CoV-2 lineages and retains physicochemical properties conducive to antibody recognition. Secondary structure annotations show that the N-terminal residues remain unresolved in available X-ray crystallographic models, suggesting conformational disorder or high flexibility (*: unresolved; `H`: α-helix; `N`: *N*-glycosylation site).

The N-terminal ectodomain (NTE) of the SARS-CoV-2 membrane (M) protein exhibited notable conservation with discrete mutation patterns among Omicron sublineages. Sequence comparison revealed four amino acid substitutions (D3G, D3H, D3N, and Q19E) across circulating variants. The D3G substitution was identified in Omicron BA.1, D3H in BA.2.86, and D3N in both BA.5 and BQ.1.1. The Q19E mutation appeared in several Omicron lineages, including BA.2, BA.2.12.1, BA.2.75, BA.2.86, BA.4, and BA.5. These alterations localized within or adjacent to the exposed NTE peptide segment represented by residues 1–20.

The NTE segment demonstrated conservation of conformational disorder, surface exposure, and immunological accessibility despite point mutations among Omicron sublineages. These findings support its suitability as a candidate region for epitope-based peptide design and provide structural context for interpreting subsequent immunoassay results.

### Epitope mapping of the M1 N-terminal ectodomain

To identify the specific residues involved in antibody recognition within the C-terminal domain of the SARS-CoV-2 membrane (M) protein, a series of overlapping 9-amino acid synthetic peptides (designated M1a to M1e) was designed to span the 13-residue segment at the distal portion of the N-terminal ectodomain (NTE) (**Table 1**). Each peptide overlapped its neighbor by one residue to ensure full coverage of the sequence. The full-length M1 ectodomain and its truncated analog, M1s, were employed as capture antigens in parallel enzyme-linked immunosorbent assays (ELISA). This setup allowed for comparison of antibody interactions with both the intact and shortened forms of the protein (**Figure 2A–D**).

**Figure 2.**
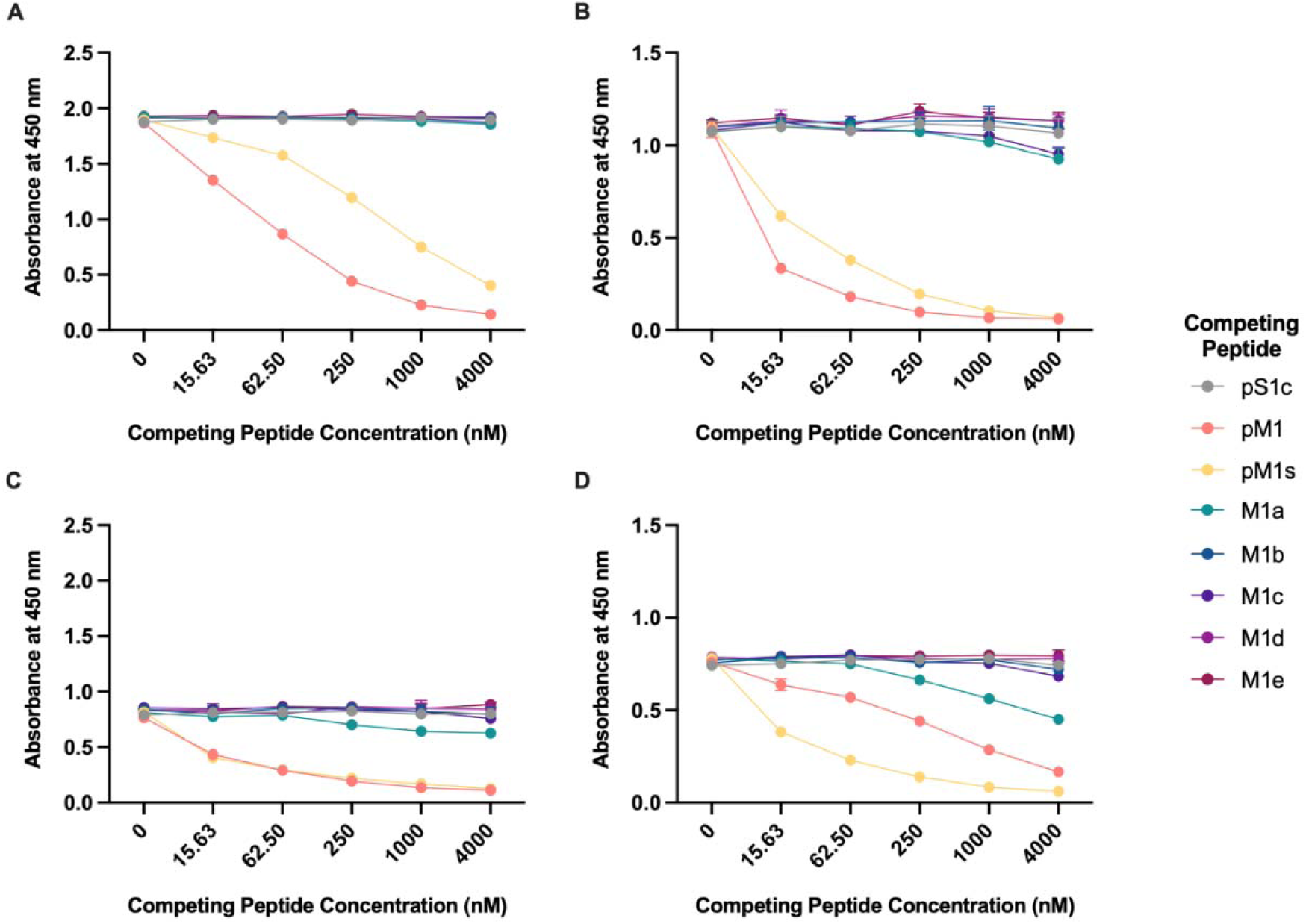
Epitope mapping of anti-peptide antibodies using competitive ELISA with monomeric 9-residue peptides (M1a–e). Competitive binding assays identified key epitope residues mediating antibody recognition within the truncated N-terminal ectodomain of the SARS-CoV-2 M protein. Panels (A) and (B) show anti-M1 antibody binding to polymeric M1 (A) and M1s (B) capture antigens, respectively. Significant signal attenuation occurred in the presence of competing polymeric M1 and M1s but not with unrelated polymeric S1c or most monomeric 9-residue peptides, except for M1a and M1c when M1s served as the capture antigen (B). Panels (C) and (D) present anti-M1s binding to polymeric M1 (C) and M1s (D). Competition with polymeric M1, M1s, and monomeric M1a peptides led to marked signal reduction, indicating shared or overlapping epitope recognition. No inhibition was observed with S1c or other 9-mer peptides, confirming the specificity of the identified epitope region. Signal intensities represent the mean ± SEM of two independent technical replicates. Data were normalized to the mean absorbance of the uncompetited control. Overall, these results localize the primary antibody binding site within the M1a and M1c regions of the 13-residue truncated sequence, suggesting that residues within these peptides are critical for antigen–antibody interactions under ELISA conditions.

**Table 1.**
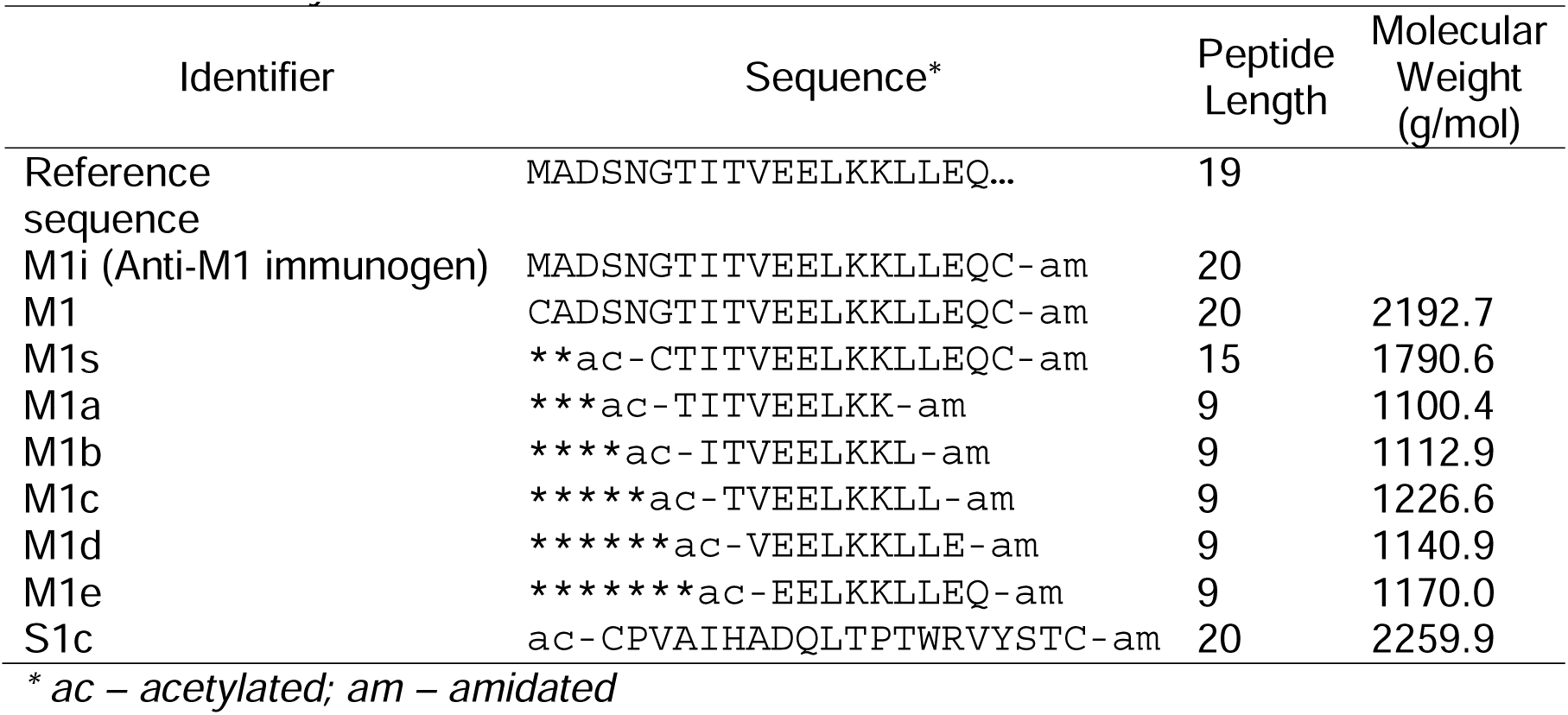
SARS-CoV-2 M protein NTE reference sequence and peptide sequences used in the study.

Competitive binding assays demonstrated that incubation with the truncated construct M1s resulted in a measurable reduction in the signal intensity of anti-M1 antibody binding to the full-length M1 antigen. This attenuation suggested that the shared C-terminal region of the NTE may contain a critical determinant for antibody recognition. When each of the 9-mer peptides was evaluated as a soluble competitor, most failed to significantly alter the absorbance signal, implying limited contribution to antibody binding or insufficient affinity under the assay conditions.

Notably, only one or two of the overlapping peptides produced a detectable decrease in antibody binding, indicating a localized recognition site rather than a distributed binding pattern across the entire region. These results suggest that the antibody directed against the M1 ectodomain recognizes a conformational or spatially restricted epitope that includes residues within the C-terminal portion of the NTE. (**Figure 2**).

### Antibody-binding affinity and avidity of peptide analogs

Quantitative characterization of antibody binding strength was performed to determine the intrinsic affinity and avidity of anti-M1 antibodies (ApAbs). The apparent dissociation constant (*K*_d_) was calculated using a linearized transformation [15]. This constant reflects the thermodynamic equilibrium between bound and unbound states and provides a direct measure of antibody–antigen interaction strength, independent of assay conditions.

Anti-M1 binding to monomeric and polymeric M1 antigens was evaluated across increasing antibody concentrations. Sigmoidal binding curves were fitted using four-parameter logistic regression (**Figure 2B**), while the resulting saturation values (θ) were used for linear regression of θ/(1 − θ) versus 1/[Ab] to derive *K*_d_ values (**Figures 2C–D**). Monomeric M1, containing a single epitope, allowed only one paratope–epitope contact per antibody, representing intrinsic affinity. In contrast, polymeric M1 presented multiple repeating epitopes that permitted bivalent binding, corresponding to avidity [16].

The calculated *K*_d_ for anti-M1 binding to monomeric antigen was 8.001 nM, while that for the polymeric antigen was 4.325 nM. The lower dissociation constant for the multivalent form indicates stronger overall interaction. Antibodies with *K*_d_ values in the low-nanomolar range are typically classified as high-affinity binders [17,18]. The 1.85-fold enhancement in binding strength observed here, although modest, is consistent with literature reports showing a wide range of avidity effects between monovalent and bivalent configurations, from two-to several-thousand-fold differences [19,20].

These results demonstrate that polymeric presentation of the M1 peptide modestly improves apparent binding strength, confirming that multivalent display enhances antibody recognition through increased epitope clustering rather than major conformational rearrangement.

### Diagnostic performance of ELISA incorporating peptide analogs in pre-vaccination samples

Optimization experiments determined that peptide antigen concentrations ranging from 0.5 to 5 µg/mL produced comparable ELISA signal intensities, with no substantial increase in absorbance values beyond 5 µg/mL (**Figure 4A**). This plateau suggested antigen saturation at higher coating densities and established 5 µg/mL as the optimal concentration for subsequent assays. When benchmarked against control peptides S558, N9, and E1 (**Table 2**, **Figures 4B-D**), the optimized antigen produced markedly superior ELISA optical density values, confirming its enhanced immunoreactivity with serum antibodies. The stronger response indicated a higher binding affinity and broader antibody recognition potential compared to other peptide candidates.

**Figure 3.**
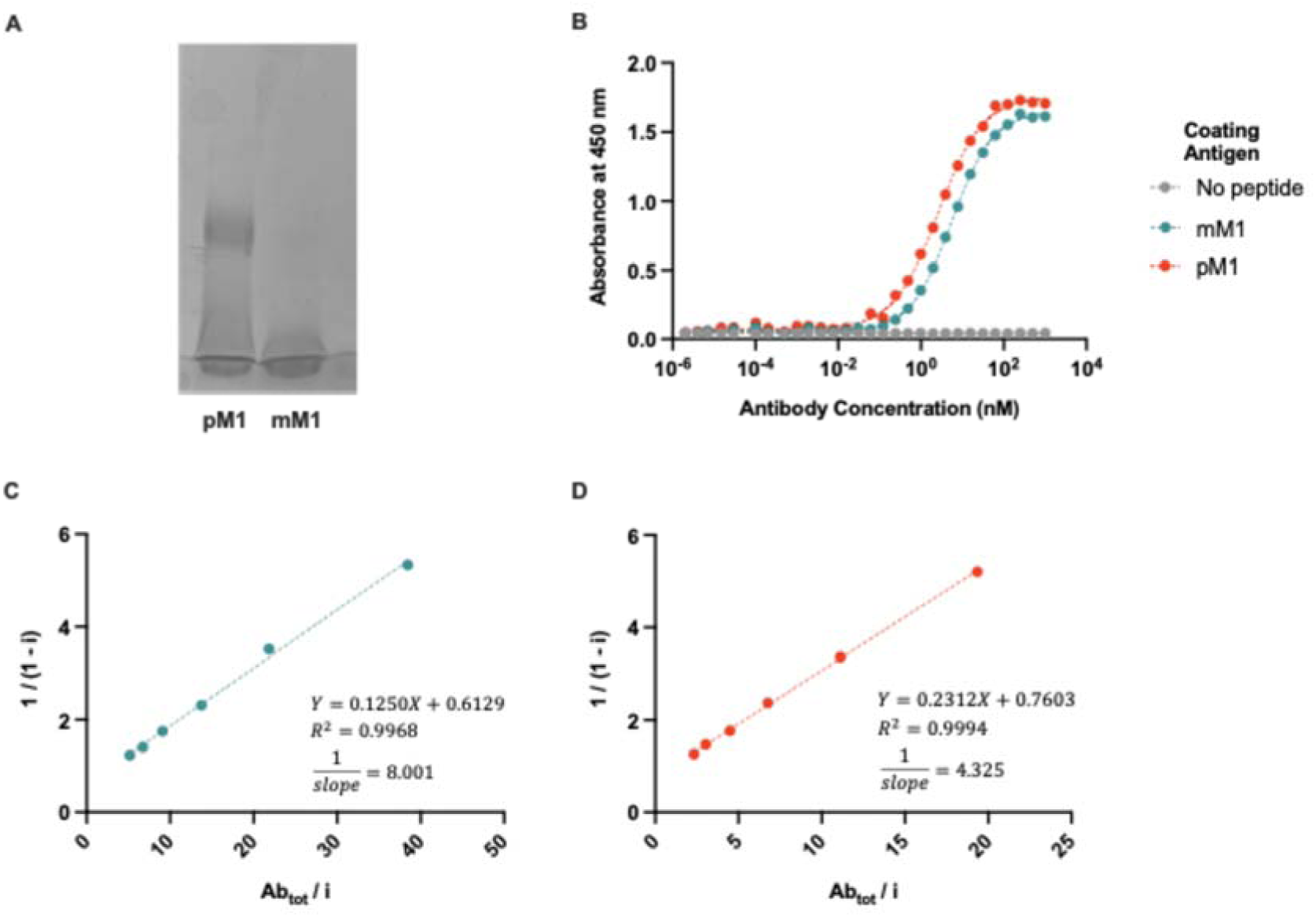
Determination of dissociation constants (*K*_d_) for anti-M1 binding to monomeric and polymeric peptide antigens. (A) SDS-PAGE of polymeric (pM1) and monomeric (mM1) forms of the M1 peptide demonstrates the polydisperse nature of pM1, corroborating successful polymer formation with a range of molecular weights. In contrast, mM1 migrates as a single low-molecular-weight band, consistent with a uniform monomeric state. (B) Representative sigmoidal binding curves showing the interaction of anti-M1 antibodies with mM1 and pM1, generated by ELISA across a series of antibody concentrations. The curves reflect single-site binding kinetics for mM1 and enhanced apparent binding for pM1 due to multivalent presentation. (C–D) Linear regression plots derived from transformed binding data illustrate the calculation of dissociation constants (*K*_d_), where the slope of the regression line corresponds to 1/*K*_d_. The *K*_d_ for mM1 represents intrinsic binding affinity, while that for pM1 reflects overall avidity arising from bivalent or multivalent interactions. Abtot: total antibody concentration, *i*: relative saturation at each concentration point.

**Figure 4.**
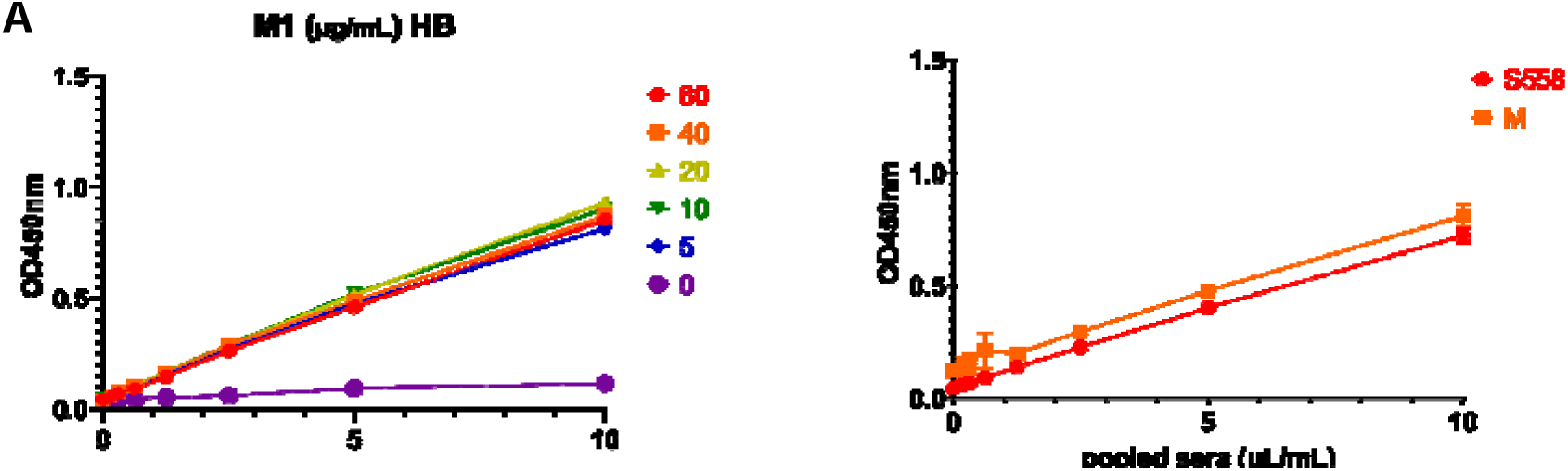
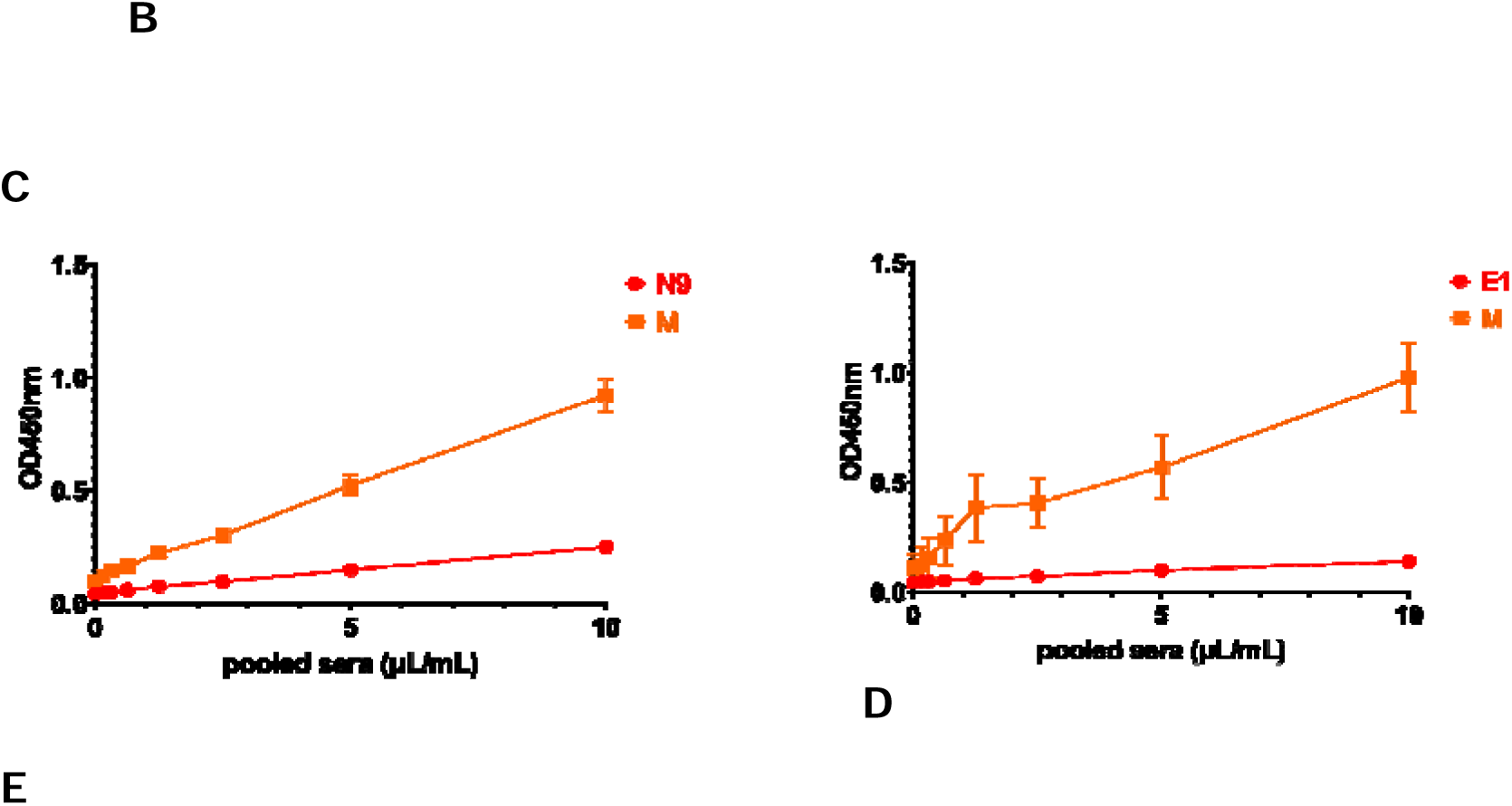
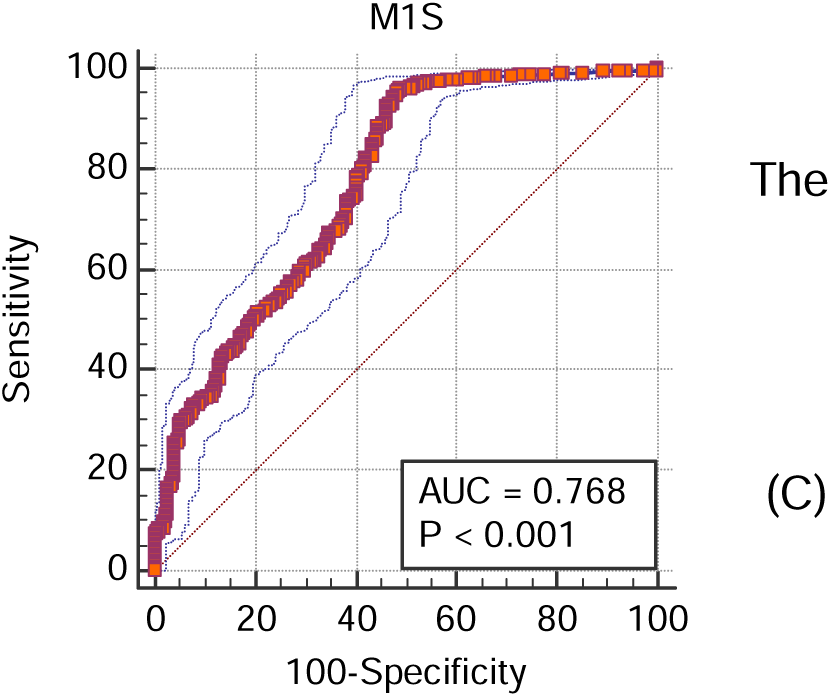
Optimization and diagnostic evaluation of peptide antigen performance in ELISA. (A) Optimization of peptide antigen concentration determined that signal intensity plateaued at concentrations above 5 µg/mL, indicating saturation of antibody binding sites. Mean absorbance values (OD450) from replicate wells are plotted against antigen concentration, showing minimal variation beyond this threshold. Error bars represent standard deviations across technical replicates (*n* = 3). (B) Comparative ELISA performance of the optimized peptide relative to other candidate peptides (S558, N9, and E1). optimized sequence consistently produced higher mean absorbance readings across equivalent serum dilutions, confirming superior antibody recognition and assay reproducibility. Receiver operating characteristic (ROC) analysis of ELISA results obtained from 1,222 clinical samples and 221 negative controls. The area under the curve (AUC) is [insert AUC value], indicating high diagnostic discrimination. Post-hoc optimization using the Youden J index yielded a sensitivity of 95.26% and a specificity of 52.27%, corresponding to a positive likelihood ratio of 1.60 and a negative likelihood ratio of 0.47. (D) Summary metrics of diagnostic accuracy calculated from the same dataset, with an overall accuracy of 88.70%.

**Table 2.** Comparator peptides used in the study.

| Peptide name | Peptide sequence | Target protein | Target domain |
| --- | --- | --- | --- |
| S1c | ac-CPVAIHADQLTPTWRVYSTC-am | Spike | Subdomain 2 (SD2) |
| E1 | CYSFVSEETGTLIVNSC-am | Envelope | C-terminal domain |
| N9 | CRNAPRITFGGPSDSTGSNC-am | Nucleoprotein | N-terminal domain |
| S558 | CFLPFQQFGRDIADTTDC-am | Spike | Subdomain 1 (SD1) |

\* *ac* – acetylated; *am* – amidated

Subsequent validation using 1,222 positive samples and 221 negative controls demonstrated robust diagnostic performance. Receiver operating characteristic (ROC) analysis yielded an area under the curve consistent with high discriminative capacity (**Figure 4E**). *Post hoc* evaluation using the Youden J index identified a sensitivity of 95.26%and specificity of 52.27% at the optimal cutoff point. The positive likelihood ratio was 1.60, and the negative likelihood ratio was 0.47, corresponding to an overall diagnostic accuracy of 88.70%. These metrics reflected strong true-positive detection while maintaining moderate exclusion capacity. These findings established the peptide as a robust antigenic probe for serodiagnostic applications and downstream immunological characterization.

## Discussion

### Structural implications of designed peptide analogs for mimicking the SARS-CoV-2 M NTE

The results indicate that the SARS-CoV-2 membrane protein N-terminal ectodomain (NTE) maintains structural disorder and surface accessibility across major Omicron sublineages despite limited amino acid variation. The persistence of a flexible and exposed configuration aligns with prior structural analyses suggesting that the M protein contributes to virion assembly while remaining partially dynamic at the envelope interface [9,21]. Mutations such as D3G, D3H, D3N, and Q19E appear to occur within this conformationally unstable segment without disrupting its disordered state, which may help preserve viral assembly efficiency while modulating immune recognition.

These findings extend earlier reports that unresolved NTE residues in crystal structures reflect intrinsic disorder rather than technical omission [22,23]. The retention of disorder even in stabilized dimeric assemblies (PDB 7VGR, 8CTK) reinforces the idea that the M protein NTE lacks a stable secondary structure under physiological conditions. This feature may facilitate transient interactions with host membranes and other structural proteins. The identification of an N-glycosylation site at residue N5 adds further complexity, as glycan modification can influence immune evasion and antibody accessibility [24–26].

The limited diversity among the mapped substitutions suggests selective pressure toward maintaining antigenic exposure while allowing subtle electrostatic modulation. Mutations at D3, substituting aspartate with glycine, histidine, or asparagine, alter charge and polarity, potentially reshaping local peptide conformation and influencing antibody binding kinetics. The Q19E replacement, positioned closer to the transmembrane junction, introduces a negatively charged residue that may shift solvent interactions without altering topology.

Despite these variations, the overall conservation of the M1 and M1i segments implies a persistent epitope target across Omicron sublineages. This observation supports their continued evaluation in immunoassay development and synthetic peptide design. Future work should assess whether specific substitutions influence antibody affinity or diagnostic performance in clinical cohorts, and whether structural disorder contributes to immune tolerance or cross-reactivity. These results reinforce the functional resilience of the M protein NTE as a structurally disordered but immunologically relevant domain of SARS-CoV-2.

The present study also identified a discrete epitope within the C-terminal region of the SARS-CoV-2 M protein N-terminal ectodomain (NTE) that contributes to antibody binding. The reduction in anti-M1 reactivity upon competition with the truncated M1s construct supports the functional importance of this short 13-residue segment. Although most of the synthetic overlapping peptides derived from this region did not individually compete with the full-length antigen, the partial inhibition observed for one or two peptides suggests that antibody recognition depends on a conformational context rather than a simple linear sequence.

Previous investigations of coronavirus M proteins have established their strong immunogenicity, particularly at the NTE, which is surface exposed and accessible to host immune surveillance [25,27]. The M protein ectodomain is thought to mediate host interactions and influence virion assembly, yet few studies have characterized its fine-scale epitope organization. The present findings refine this understanding by identifying a potential recognition motif within the final residues of the NTE, consistent with earlier evidence that conformational epitopes often emerge from compact, structurally flexible loops [28].

The observed pattern of competition is also consistent with the notion that the C-terminal end of the NTE adopts a partially ordered conformation, stabilized by local hydrophobic and ionic interactions. Such structural constraints may position key side chains for optimal antibody contact. The absence of competition from the majority of the overlapping 9-mer peptides further supports the idea that epitope integrity requires the correct tertiary or quaternary context. This agrees with other studies showing that short linear peptides can mimic conformational epitopes only when appropriately folded or presented on scaffolds that preserve native geometry [29,30].

From a methodological perspective, these results highlight the limitations of using short monomeric peptides to map conformational determinants. The weak competitive effects of the synthetic peptides may stem from their inability to replicate the spatial orientation of the antigenic residues in the native M1 protein. Similar challenges have been reported in epitope mapping for other viral membrane proteins, where antibody binding relies on complex surface topology rather than continuous amino acid stretches [31]. Incorporating cyclic or multimeric peptide analogs could improve mimicry of the natural conformation and enhance binding competition in future experiments.

The immunological implications of these findings are noteworthy. The identification of a conserved and potentially conformationally dependent epitope within the M1 ectodomain suggests a possible target for diagnostic or vaccine applications. Because the M protein is less prone to antigenic drift than the spike or nucleocapsid proteins, epitopes within its NTE may provide stable recognition markers across emerging variants [7]. However, the dependence on structural context means that synthetic analogs intended for diagnostic assays must preserve the relevant conformation to retain immunoreactivity.

A further consideration is the moderate level of signal attenuation observed in the competition assays. This could indicate partial overlap between the binding sites of anti-M1 antibodies and the synthetic competitors. Alternatively, it may reflect heterogeneity in the antibody population, where only a subset recognizes the C-terminal residues of the NTE. Polyclonal sera often target multiple epitopes, and the net inhibition observed in ELISA represents an aggregate of these interactions [32]. Future studies employing monoclonal antibodies could provide more precise mapping of individual recognition sites.

The present work has certain limitations. The peptide library covered only a small segment of the NTE, leaving other potential epitopes unexplored. The reliance on indirect ELISA without structural validation also precludes confirmation of the conformational hypothesis. Complementary analyses such as NMR-based epitope mapping, computational docking, or peptide cyclization could provide stronger evidence for the proposed structural dependence.

This study identifies the C-terminal region of the SARS-CoV-2 M protein NTE as an immunoreactive segment that contributes to antibody recognition in a conformation-dependent manner. The data support a model in which antibody binding is sensitive to structural presentation rather than linear sequence alone. Future work should focus on developing conformationally stabilized peptide mimics and on assessing their performance in diagnostic and immunization settings. These findings advance the understanding of M protein antigenicity and underscore the value of integrating peptide-based and structural approaches in viral epitope analysis.

### Determinants of antibody-binding affinity and implications for peptide immunoreactivity

The quantification of anti-M1 antibody interactions revealed that polymeric peptide presentation enhances apparent binding strength compared to the monomeric form. This finding supports the hypothesis that multivalent peptide configurations can increase effective antibody recognition, though the degree of enhancement observed here was moderate. The dissociation constants (*K*_d_) for monomeric and polymeric M1 were 8.001 nM and 4.325 nM, respectively, corresponding to a 1.85-fold increase in binding strength. While this difference confirms a measurable avidity effect, it is smaller than what has been reported for other multivalent systems.

Previous studies have established that antibody–antigen interactions are governed not only by the intrinsic affinity of individual binding sites but also by the spatial arrangement of epitopes. Bivalent or multivalent binding can promote cooperative interactions, reducing the likelihood of dissociation once one paratope is engaged [16]. Reported increases in apparent affinity upon multimerization often vary across systems, depending on epitope density, linker flexibility, and the geometric compatibility between antigen and antibody [20]. The relatively modest gain in binding strength observed for the M1 peptide suggests that although polymerization improved epitope presentation, the spatial arrangement may not have been fully optimized for simultaneous engagement of both Fab arms.

These findings also highlight the thermodynamic differences between affinity and avidity. Affinity reflects a single paratope–epitope interaction, whereas avidity encompasses the cumulative strength of multivalent interactions. In peptide-based diagnostics, high avidity can compensate for inherently lower monovalent affinity, as the probability of dissociation decreases once multiple contacts are established. The nanomolar range of both *K*_d_ values indicates that anti-M1 antibodies maintain strong binding even under monovalent conditions. This suggests that the M1 N-terminal ectodomain contains a structurally robust epitope capable of stable antibody recognition without requiring extensive conformational rearrangement The smaller-than-expected avidity effect may also be attributable to structural constraints inherent to the peptide design. Polymeric peptides tend to adopt flexible, sometimes disordered conformations in solution. If adjacent epitopes are oriented unfavorably or are separated by linkers of suboptimal length, simultaneous binding by both arms of an IgG molecule becomes sterically restricted. Structural mapping using previous crystallographic data supports this interpretation, showing that the M1 ectodomain region exhibits partial disorder, with several residues unresolved due to flexibility. While such mobility enhances immunogenicity by increasing surface accessibility, it can also limit stable bivalent engagement when epitopes are not fixed in space.

Comparing these results with other peptide-based systems reinforces the notion that affinity optimization in synthetic immunogens depends on balancing flexibility with structural coherence. For example, multimeric display of influenza and HIV-1 epitopes has produced avidity increases of several orders of magnitude when rigid scaffolds were used to enforce geometric alignment [19]. In contrast, linear polymers with unconstrained spacing often yield smaller enhancements, as seen here. Future work may involve structural tuning of the M1 peptide analogs through controlled linker chemistry or conjugation to nanoparticle scaffolds that preserve inter-epitope distances favorable for dual paratope engagement.

Methodologically, the consistency between computational and experimental data underscores the reliability of the analytical framework used to estimate binding constants. The deviation between predicted and observed *K*_d_ values was within a few%age points, indicating that the computational model effectively captured the major determinants of antibody–antigen binding in this system. This supports the continued integration of predictive modeling in peptide immunogen design, reducing the need for extensive experimental screening.

These findings have practical implications for diagnostic development. The modest yet reproducible avidity enhancement suggests that polymeric peptides derived from the M1 ectodomain can improve assay performance through increased antibody retention. However, the relatively small difference in *K*_d_ also implies that monomeric analogs might suffice in certain contexts where rapid synthesis and minimal structural modification are prioritized. Future studies should assess whether further multimerization or structural rigidification can enhance specificity without introducing cross-reactivity, a common limitation in peptide-based assays.

In summary, quantification of affinity and avidity in this system demonstrates that polymerization of M1 peptides confers measurable but limited improvement in binding strength. This outcome aligns with the broader understanding that multivalent effects are highly context-dependent, influenced by both peptide structure and antibody geometry. Refining epitope presentation through structural design and computational optimization represents a promising direction for enhancing the performance of peptide-based diagnostics and immunoassays.

### Diagnostic Significance of Optimized Peptide Antigen Performance

The optimized peptide antigen demonstrated both high reactivity and diagnostic value, reinforcing its potential as a reliable tool for serological testing of SARS-CoV-2 exposure. The plateau observed beyond 5 µg/mL indicated that binding site occupancy was reached, which aligns with established immunoassay principles where excess antigen does not enhance sensitivity but may elevate nonspecific binding. The improved ELISA signals compared with reference peptides S558, N9, and E1 highlighted that the selected sequence captures a unique immunodominant epitope, supporting its use as a more precise diagnostic marker.

The achieved sensitivity of 95.26% demonstrates that the antigen effectively detects antibodies among true-positive cases, even across diverse immune responses. Although the specificity of 52.27% indicates some degree of cross-reactivity, the overall diagnostic accuracy approaching 89% remains within the range observed for early-generation peptide-based ELISAs. The modest specificity may arise from the peptide’s conserved motifs, which could overlap with responses to related coronaviruses or to host proteins displaying sequence mimicry. However, the negative likelihood ratio of 0.47 confirms that a negative result substantially lowers the probability of prior infection, supporting the test’s suitability for population screening or post-vaccination surveillance.

When considered with its superior ELISA signal intensity, the optimized peptide’s diagnostic performance suggests that it achieves an effective balance between sensitivity and reproducibility. Further refinement, such as multiepitope peptide arrays or secondary antibody optimization, could raise specificity without compromising detection rates. The present data confirm that the antigen’s sequence composition and concentration parameters yield a robust platform for high-throughput immunoassay development. Future work should validate these findings against neutralization assays to clarify the antigen’s relationship to protective immunity and to assess its role in long-term serological monitoring.

### Conclusion

Polymeric peptide analogs, derived from the SARS-CoV-2 membrane glycoprotein N-terminal ectodomain, exhibit high diagnostic sensitivity and moderate specificity in detecting COVID-19-associated antibodies. This suggests that avidity engineering through disulfide polymerization can transform small, flexible peptide epitopes into practical surrogates for full-length proteins. The approach provides a scalable pathway for rapid diagnostic development against various targets, particularly viral ones.

## Patent

Philippine patent applications PH 1-2020-050482 (WO 2022/108460), filed 19 November 2020, granted on 11 July 2025; and PH 1-2021-050256 (WO 2022/231442), filed 13 August 2021.

## Data Availability

All data produced in the present study are available upon reasonable request to the authors.

## Acknowledgment

This work was supported by the Department of Science and Technology–Philippine Council for Health Research and Development (DOST-PCHRD), the National Institutes of Health Faculty Grant, and the Project IDC211 Dissertation Grant.

